# Does engagement in HIV care affect screening, diagnosis, and control of noncommunicable diseases in sub-Saharan Africa? A systematic review and meta-analysis

**DOI:** 10.1101/2023.01.30.23285196

**Authors:** Emma M Kileel, Amy Zheng, Jacob Bor, Matthew P Fox, Nigel J Crowther, Jaya A George, Siyabonga Khoza, Sydney Rosen, Willem DF Venter, Frederick Raal, Patricia Hibberd, Alana T Brennan

**Author notes:** Corresponding Author: Emma M Kileel, 715 Albany Street, T3/5E Boston, MA 02118. **Conflicts of Interest and Sources of Funding**. This work was supported by National Institute of Diabetes and Digestive and Kidney Diseases 1K01DK116929-01A1 (ATB and EMK) and the Bill & Melinda Gates Foundation INV-031690 (SR). For the remaining authors none were declared.

## Abstract

**Objective:** Low- and middle-income countries are facing a growing burden of noncommunicable diseases (NCDs). Providing HIV treatment may also provide opportunities to increase access to NCD services in under-resourced environments. We sought to investigate whether reported use of antiretroviral therapy (ART) was associated with increased screening, diagnosis, treatment, and/or control of diabetes, hypertension, chronic kidney disease, or cardiovascular disease among people living with HIV (PLWH) in sub-Saharan Africa (SSA).

**Design:** Systematic review and meta-analysis.

**Methods:** We searched 10 electronic literature databases for studies published between 01 January 2011 and 31 December 2022 using a comprehensive search strategy. We sought studies reporting on screening, diagnosis, treatment, and/or control of NCDs of interest by ART use among non-pregnant adults with HIV ≥16 years of age in SSA. Random effects models were used to calculate summary odds ratios (ORs) of the risk of diagnosis by ART status and corresponding 95% confidence intervals (95% CIs), where appropriate.

**Results:** Twenty-six studies, describing 13,570 PLWH in SSA, 61% of whom were receiving ART, were included. ART use was associated with a small but imprecise increase in the odds of diabetes diagnosis (OR: 1.07; 95% CI: 0.71, 1.60) and an increase in the odds of hypertension diagnosis (OR: 2.10, 95% CI: 1.42, 3.09). We found minimal data on the association between ART use and screening, treatment, or control of NCDs.

**Conclusion:** Despite a potentially higher NCD risk among PLWH and regional efforts to integrate NCD and HIV care, evidence to support effective care integration models is lacking.

## Introduction

The prevalence of noncommunicable diseases (NCDs) is on the rise globally [1], [2]. In low- and middle-income countries (LMICs), the disease burden has already shifted from predominantly infectious diseases to predominantly NCDs [3], [4]. By 2030, NCDs are expected to account for nearly 46% of deaths in sub-Saharan Africa (SSA), compared to 28% in 2008 [5]–[7]. In SSA where there is already a heavy burden of HIV and other communicable diseases, this creates a challenging multimorbidity disease environment. Co-existence of HIV with one or more NCDs, such as type 2 diabetes mellitus (diabetes) or hypertension, increases the complexity of patients’ disease and care management profile, contributing to poorer health outcomes and increased health care costs for both the individual and the country [8]–[10].

People living with HIV (PLWH) may be at increased risk for NCDs. Antiretroviral therapy (ART) has resulted in a decrease in HIV infections and an increase in the lifespan of PLWH [11], [12]. However, chronic inflammation as a result of long-term HIV infection is associated with an increased risk of chronic conditions, including chronic kidney disease (CKD) and cardiovascular disease (CVD) [13],[14]. Moreover, emerging data have demonstrated substantial weight gain associated with newer ART regimens, such as integrase-strand-inhibitors [15]–[17]. As overweight and obesity are known risk factors for numerous NCDs, including hypertension, diabetes, and CVD [18], [19], this ART-associated weight gain could result in an increase of NCD rates in this population.

Though HIV infection and ART use may contribute to increases in NCD risk, the HIV care structure may provide a gateway to improve rates of detection and treatment of NCDs among PLWH. Routine NCD screening among healthy, asymptomatic adults in SSA is not common [20]. Traditionally, however, PLWH on ART have been required to present to a clinic monthly for the first 6 months after ART initiation and then at least every 3 or 6 months once stable in order to obtain their medications [21]. ART guidelines typically include collection of data on blood glucose and urine protein at treatment initiation as well as routine collection of data on weight and blood pressure [22]–[24]. This increased contact with the healthcare system could increase the likelihood of NCD diagnosis, treatment, and control among PLWH. The purpose of this report was to investigate whether reported use of ART was associated with increased screening, diagnosis, treatment, and/or control of four common NCDs – diabetes, hypertension, CKD, and CVD – among PLWH in SSA, a region with an estimated 28.7 million people prescribed ART and engaged in care [25].

## Methods

### Search strategy and selection criteria

The protocol for this meta-analysis was registered with PROSPERO (CRD42021290283). To conduct the systematic review, we ran comprehensive searches on 3 electronic literature databases (PubMed, EMBASE, Web of Science) and Conference on Retroviruses and Opportunistic Infections (CROI), International AIDS Society (IAS), International Diabetes Federation World Congress, American Diabetes Association, American Heart Association Scientific Sessions, Hypertension Scientific Sessions, Kidney Week abstract archives. Articles and abstracts published between 01 January 2011 and 31 December 2022 that reported outcomes among non-pregnant adults ≥ 16 years of age with HIV in SSA were included for review. We further applied snowball sampling methods to articles selected for review [26].

Titles and abstracts of search results were imported into Covidence systematic review software [27] and screened independently by two reviewers (E.M.K and A.Z). A senior reviewer (A.B.) resolved any discrepancies. Titles and abstracts that contained potentially relevant information were identified and selected for full-text review. Articles or abstracts that reported on the prevalence of screening, diagnosis, or treatment by ART status of a least one disease of interest and did not meet any exclusion criteria (Supplemental Table 2), were selected for data extraction. Quality of studies was assessed using The Joanna Briggs Institute Critical Appraisal Tools for analytical cross-sectional studies [28]. Eight questions were used to evaluate the quality of each individual study, with the options of “yes”, “no”, “unclear”, or “not applicable”. We assigned values to the response options to obtain an overall quality score.

### Exposure

The exposure of interest in this review was reported ART use (versus no reported ART use) among non-pregnant adults with HIV in SSA.

### Outcomes

Outcomes of interest were:

1) self-reported **screening** or documentation of laboratory test for diabetes, hypertension, CKD, or cardiovascular disease;
2) self-reported or clinical **diagnosis** based on study criteria of diabetes, hypertension, CKD, or cardiovascular disease;
3) reported (via self-report or documentation in medical records) use of **treatment** for diabetes, hypertension, CKD, or cardiovascular disease;
4) reported **control** of diabetes or hypertension based on clinical or lab-defined criteria or reported management of CKD or cardiovascular disease.

Study specific definitions of NCD diagnosis are included in Table 1. If studies reported an outcome by multiple indicators, the method with the largest sample size was included in the meta-analysis. Study specific definitions of treatment, screening, or control of NCDs are included in Supplemental Table 3.

**Table 1.** Characteristics of included studies grouped by noncommunicable disease (NCD) and odds of NCD diagnosis comparing HIV-positive ART exposed to HIV-positive ART unexposed in each study.

| Study No | Study, Year Published | Country | Dates of observation | Total N (% ART+) | Total female, n (%) | Mean Age (years) | Screened ART+, n (%) | Screened ART-, n (%) | Diagnostic Criteria | Diagnosed ART+, n (%) | Diagnosed ART-, n (%) | Initiated Treatment ART+, n (%) | Initiated Treatment ART-, n (%) | Control among treated ART+, n (%) | Control among treated ART-, n (%) | Odds ratio of NCD diagnosis<sup>1</sup> (95% CI) |
| --- | --- | --- | --- | --- | --- | --- | --- | --- | --- | --- | --- | --- | --- | --- | --- | --- |
| Outcome=Diabetes |  |  |  |  |  |  |  |  |  |  |  |  |  |  |  |  |
| 1 | Dave et al, 2011 [47] | South Africa | | 849 (52) | 653 (77) | | | | Fasting glucose ≥ 7 mmol/L or or OGTT ≥ 11.1 mmol/L | 10 (2.2) | 14 (3.4) | | | | | 0.65 (0.28, 1.47) |
| 2 | Sani et al, 2013 [48] | Nigeria | 05/09 - 08/09 | 200 (50) | 106 (53) | 32.5 | | | Fasting glucose ≥ 7 mmol/L or Random glucose ≥ 11.1 mmol/L | 3 (3) | 3 (3) | | | | | 1.0 (0.20, 5.08) |
| 3 | Peck et al, 2014 [49] | Tanzania | 10/12 - 04/13 | 301 (50) | 204 (68) | 38.5<sup>4</sup> |  |  | Self-reported via WHO STEPS questionnaire | 1 (0.7) | 1 (0.7) |  |  |  |  | 1.01 (0.06, 16.25) |
| 4 | Shey Nsagha et al, 2015 [50] | Cameroon | 03/14 - 08/14 | 215 (74) | 161 (75) | 43.2 | | | Fasting glucose ≥ 7 mmol/L | 3 (1.9) | 2 (3.6) | | | | | 0.51 (0.08, 3.11) |
| 5 | Osegbe et al, 2016 [51] | Nigeria | | 200 (50) | | 36.6 | | | Fasting glucose ≥ 7 mmol/L | 9 (9) | 2 (2) | | | | | 4.85 (1.02, 23.03) |
| 6 | Divala et al, 2016 [52] | Malawi | 7/14 - 10/14 | 952 (96) | 683 (72) | 43.0 | | | Elevated glucose defined as fasting glucose ≥ 7 mmol/L or random glucose ≥ 11.1 mmol/L on two occasions or reported use of antidiabetic drugs | 38 (4.2) | 1 (2.6) | | | | | 0.61 (0.08, 4.53)<sup>2</sup> |
| 7 | Kingery et al, 2016 [53] | Tanzania | 10/12 - 4/13 | 301 (50) | 204 (68) | 40.5 | | | Fasting glucose ≥ 7 mmol/L or Random glucose ≥ 11.1 mmol/L | 27 (18) | 1 (0.7) | | | | | 32.93 (4.41, 245.78) |
| 8 | Manne-Goehler et al, 2017 [39] | South Africa | 11/14 - 11/15 | 1035 (64) | 560 (54) | 55.4 | 324 (49) | 157 (42) | Self-reported diabetes diagnosis or glucose ≥ 7 mmol/L) in fasting group (defined as > 8 hours), glucose ≥11.1 mmol/L in nonfasting (“random or casual”) samples. Individuals with missing fasting information were considered to be not fasting. | 52 (7.8) | 23 (6.3) | 363 (55) | 162 (44) | | | 1.30 (0.78, 2.16) |
| 9 | Osoti et al, 2018 [54] | Western Kenya | 07/14 - 09/14 | 300 (55) | 192 (64) | 40.7 | | | Fasting blood glucose ≥ 7 mmol/L | 2 (1.5) | 5 (3.1) | | | | | 0.32 (0.06, 1.69) |
| 10 | Achwoka et al, 2019 [32] | Kenya | 10/15 - 09/16 | 3170 (68) | 2115 (67) |  |  |  | Diabetes mellitus documented in medical records | 7 (0.3) | 2 (0.2) |  |  |  |  | 1.61 (0.33, 7.79) |
| 11 | Manne-Goehler et al, 2019 [55] | South Africa | 11/14 - 11/15 | 1035 (71) | 560 (54) | 55.4 | | | Fasting glucose ≥ 7 mmol/L or Random glucose ≥ 11.1 mmol/L or self-reported diagnosis of diabetes that had been made by a doctor, nurse, or healthcare worker, or self-reported use of medication for diabetes prescribed by a doctor, nurse, or healthcare worker | 59 (8.0) | 21 (7.1) | | | | | 1.17 (0.69, 1.95) |
| 12 | Sarfo et al, 2019 [31] | Ghana |  | 451 (55) | 367 (81) | 44.5 |  |  | History of known diabetes, use of diabetes medication, or HbA1c > 6.5% | 21 (8.4) | 19 (9.5) |  |  |  |  | 0.88 (0.46, 1.68) |
| 13 | Jeremiah et al, 2020 [56] | Tanzania | 10/16 - 11/17 | 1292 (26) | 786 (61) | 39.9 | | | HbA1c ≥ 6.5% | 26 (7.8) | 169 (17.7) | | | | | 0.39 (0.25, 0.60) |
| | | | | | | | | | OGTT ≥ 11.1 mmol/L | 11 (3.3) | 87 (9.1) | | | | | 0.34 (0.18, 0.64) |
| | | | | | | | | | Both HbA1c ≥ 6.5% and OGTT ≥ 11.1 mmol/L | 3 (0.9) | 33 (3.5) | | | | | 0.25 (0.08, 0.83) |
| 14 | Sarfo et al, 2021 [57] | Ghana | | 502 (51) | 377 (75) | | | | History of diabetes, use of medications for diabetes, or HbA1c ≥ 6.5% | 19 (7.4) | 16 (6.6) | | | | | 1.14 (0.57, 2.27) |
| | | | | | | | | | Two fasting glucose readings ≥ 7 mmol/L | 30 (11.6) | 38 (15.6) | | | | | 1.14 (0.57, 2.27) |
| 15 | Roozen et al, 2021 [58] | South Africa | 07/16 - 11/17 | 394 (73) | 264 (67) | 39.8 |  |  | Random glucose > 11 mmol/L or use of blood glucose lowering medication | 6 (2.1) | 1 (0.9) |  |  |  |  | 2.23 (0.27, 18.78) |
| 16 | Msoka et al, 2021 [59] | Tanzania | | 104 (62) | 74 (71) | 54.6 | | | Fasting glucose≥ 7 mmol/L | 6 (9.4) | 2 (5.0) | | | | | 1.97 (0.38, 10.25) |
| Outcome=Hypertension |  |  |  |  |  |  |  |  |  |  |  |  |  |  |  |  |
| 17 | Ekali et al, 2013 [60] | Cameroon |  | 143 (80) | 103 (72) | 39.5 |  |  | Blood pressure equal to or greater than 140/90 mmHg | 18 (15.7) | 3 (10.7) |  |  |  |  | 1.55 (0.42, 5.67) |
| 2 | Sani et al, 2013 [48] | Nigeria | 05/09 - 08/09 | 200 (50) | 106 (53) | 32.5 |  |  | Blood pressure equal to or greater than 140/90 mmHg | 17 (17) | 2 (2) |  |  |  |  | 10.04 (2.25, 44. 71) |
| 18 | Botha et al, 2014 [61] | South Africa | 2010 | 137 (48) | 96 (70) | 47.7 |  |  | Self-report of previous diagnosis | 20 (31.8) | 29 (40.9) | 9 (14.0) | 11 (16.0) |  |  | 0.63 (0.31, 1.28) |
| 3 | Peck et al, 2014 [49] | Tanzania | 10/12 - 04/13 | 301 (50) | 204 (68) | 38.5<sup>4</sup> |  |  | Blood pressure equal to or greater than 140/90 mmHg | 43 (28.7) | 8 (5.3) |  |  |  |  | 7.18 (3.24, 15.91) |
| 19 | Ogunmola et al, 2014 [62] | Nigeria |  | 250 (52) | 156 (62) | 38.1 |  |  | Blood pressure equal to or greater than 140/90 mmHg | 16 (12.3) | 19 (15.8) |  |  |  |  | 1.47 (0.37, 5.84) |
| 4 | Shey Nsagha et al, 2015 [50] | Cameroon | 03/14 - 08/14 | 215 (74) | 161 (75) | 43.2 |  |  | Blood pressure equal to or greater than 140/90 mmHg | 47 (29.4) | 8 (14.5) |  |  |  |  | 2.4 (1.07, 4.52) |
| 20 | Ogunmola et al, 2015 [63] | Nigeria |  | 121 (63) | 86 (71) | 38.1 |  |  | Blood pressure equal to or greater than 140/90 mmHg | 9 (11.8) | 4 (8.9) |  |  |  |  | 1.38 (0.40, 4.76) |
| 5 | Osegbe et al, 2016 [51] | Nigeria |  | 200 (50) |  | 36.6 |  |  | SBP equal to or greater than 140 mmHg | 43 (43) | 23 (23) |  |  |  |  | 2.19 (1.02, 4.69) |
|  |  |  |  |  |  |  |  |  | DBP equal to or greater than 90 mmHg | 20 (20) | 11 (11) |  |  |  |  | 2.02 (0.91, 4.48) |
| 21 | Dimala et al, 2016 [64] | Cameroon | 04/13 - 06/13 | 200 (50) | 140 (70) | 39.1 |  |  | Blood pressure equal to or greater than 140/90 mmHg | 38 (38) | 19 (19) |  |  |  |  | 2.2 (1.07, 4.52)<sup>3</sup> |
| 6 | Divala et al, 2016 [52] | Malawi | 7/14 - 10/14 | 952 (96) | 683 (72) | 43.0 |  |  | Blood pressure equal to or greater than 140/90 mmHg or taking anti-hypertensive medication | 216 (23.7) | 10 (25.6) |  |  |  |  | 1.11 (0.53, 2.32) |
| 7 | Kingery et al, 2016 [53] | Tanzania | 10/12 - 4/13 | 301 (50) | 204 (68) | 40.5 |  |  | Blood pressure equal to or greater than 140/90 mmHg | 43 (28.7) | 8 (5.3) | 10 (6.7) | 1 (0.7) |  |  | 7.18 (3.24, 15.91) |
| 22 | Nduka et al, 2016 [65] | Nigeria | 08/14 - 11/14 | 406 (75) | 278 (69) | 37.8 |  |  | Blood pressure equal to or greater than 140/90 mmHg or current use of antihypertensive medication | 50 (16.3) | 9 (9.0) | 24 (7.8) | 4 (4.0) |  |  | 1.97 (0.93, 4.18) |
| 8 | Manne-Goehler et al, 2017 [39] | South Africa | 11/14 - 11/15 | 1035 (64) | 560 (54) | 55.4 | 473 (71.5) | 258 (69.2) | Self-report of previous diagnosis, or systolic blood pressure ≥ 140 mmHg or diastolic ≥ 90 mmHg, or self-reported use of anti-hypertensive medication at time of interview | 256 (38.7) | 162 (43.5) | 383 (57.8) | 183 (49.1) | | | 0.82 (0.63, 1.06) |
| 10 | Achwoka et al, 2019 [32] | Kenya | 10/15 – 09/16 | 3170 (68) | 2115 (67) |  |  |  | Two or more measures taken within 12 months of systolic blood pressure ≥ 140 mmHg or diastolic blood pressure ≥ 90 mmHg | 319/2170 (14.7) | 24/1000 (2.4) |  |  |  |  | 7.01 (4.60, 10.69) |
| 11 | Manne-Goehler et al, 2019 [55] | South Africa | 11/14 - 11/15 | 1035 (71) | 560 (54) | 55.4 |  |  | Self-report of ever diagnosed, or SBP ≥ 140 mmHg or DBP ≥ 90 mmHg, or self-reported use of anti-hypertensive medication at time of interview | 339 (46.2) | 143 (47.7) |  |  |  |  | 0.95 (0.72, 1.24) |
| 12 | Sarfo et al, 2019 [31] | Ghana |  | 451 (55) | 367 (81) | 44.5 |  |  | Blood pressure greater than or equal to 140/90 mmHg or history of hypertension or use of antihypertensive drugs | 92 (36.9) | 47 (23.4) | 37 (40.2) | 21 (44.7) | 11 (29.7) | 9 (42.9) | 1.91 (1.26, 2.89) |
| 15 | Roozen et al, 2021 [58] | South Africa | 07/16 - 11/17 | 394 (73) | 264 (67) | 39.8 |  |  | SBP > 130 mmHg, DBP > 85mmHg or use of antihypertensive medication | 101 (35.1) | 20 (18.9) |  |  |  |  | 2.32 (1.35, 4.00) |
| 16 | Msoka et al, 2021 [59] | Tanzania |  | 104 (62) | 74 (71) | 54.6 |  |  | Blood pressure equal to or greater than 140/90 mmHg | 14 (21.9) | 6 (15.0) |  |  |  |  | 1.59 (0.55, 4.54) |
| Outcome=Chronic Kidney Disease |  |  |  |  |  |  |  |  |  |  |  |  |  |  |  |  |
| 23 | Cailhol et al, 2011 [66] | Burundi | 2/08 - 2/09 | 238 (72) | 211 (70) | 40.1 |  |  | GFR<sub>CG</sub> < 60 ml/min or GFR<sub>MDRD</sub> < 60 ml/min/1.73m<sup>2</sup> and/or kidney damage defined as a urinary abnormality at any GFR level | 7 (4) | 8 (12) |  |  |  |  | 0.31 (0.11, 0.91) |
| 24 | Sarfo et al, 2013 [67] | Ghana | 05/10 - 01/11 | 238 (61) | 172 (72) | 39.2<sup>4</sup> |  |  | CrCl < 60 ml/min/1.73m<sup>2</sup> (Cockcroft-Gault) or proteinuria ≥ 1 (confirmed by uPCR > 45 mg/mmol) on urinalysis | 22 (15) | 41 (44) |  |  |  |  | 3.63 (1.67, 7.91) |
| 3 | Peck et al, 2014 [49] | Tanzania | 10/12 - 04/13 | 301 (50) | 204 (68) | 38.5<sup>4</sup> |  |  | eGFR <60 mL/minute and/or microalbuminuria according to KDIGO | 62 (41) | 52 (34) |  |  |  |  | 1.34 (0.84, 2.14) |
| 12 | Sarfo et al, 2019 [31] | Ghana |  | 451 (55) | 367 (81) | 44.5 |  |  | eGFR <60 mL/minute | 6 (2) | 6 (3) |  |  |  |  | 0.80 (0.25, 2.52) |
| 25 | Che Awah Nforbugwe et al, 2020 [68] | Cameroon | 3/16 - 6/16 | 136 (50) | 100 (72) | 39.5 |  |  | CrCl < 60 ml/min/1.73m<sup>2</sup> | 18 (27) | 8 (12) |  |  |  |  | 2.7 (1.08, 6.73) |
| 26 | Sulaimain et al, 2022 [69] | Nigeria | 01/12- 12/14 | 400 (50) | 276 (69) | 36.6 |  |  | eGFR <60 mL/minute | 21 (10.5) | 61 (30.5) |  |  |  |  | 0.27 (0.16, 0.46) |
| Outcome=Cardiovascular Disease |  |  |  |  |  |  |  |  |  |  |  |  |  |  |  |  |
| 10 | Achwoka et al, 2019 [32] | Kenya | 10/15 - 09/16 | 3170 (68) | 2115 (67) |  |  |  | Documented record of hypertension, heart attack, or stroke in medical records | 322 (15) | 25 (3) |  |  |  |  | 6.80 (4.49, 10.28) |

<sup>1</sup> Odds ratios represent odds of NCD diagnosis by ART status.
<sup>2</sup> Odds ratio as reported in primary paper, adjusted for age, sex, study site, and educational status.
<sup>3</sup> Odds ratio as reported in primary paper, adjusted for age, gender, family history of hypertension, smoking, and obesity.
<sup>4</sup> Median age reported
\*Abbreviations: SBP=systolic blood pressure, DBP=diastolic blood pressure, OGTT=oral glucose tolerance test, KDIGO=Kidney Disease Improvement Global Outcomes
\*Imperial to metric units conversions: 7 mmol/L = 126 mg/dL; 11.1 mmol/L = 200 mg/dL

### Data extraction

We extracted publication year, country, study design, dates of observation, mean or median age of the cohort, total number of participants, the proportion of female participants, and the number of participants who reported ART use and no ART use as reported for each study. For outcomes of interest, we extracted, if reported, the total number of participants who were screened for, diagnosed with, treated for, or achieved control levels for NCDs of interest by ART status. If reported, we extracted effect estimates – adjusted when available, otherwise crude – and 95% confidence intervals (CIs) estimating the likelihood of disease diagnosis by ART status. If odds ratios were not reported, we calculated the crude odds ratios and 95% CIs of NCD diagnosis comparing PLWH with reported ART use to PLWH reporting no ART use.

### Data analysis

We performed meta-analyses with random effects models to estimate pooled odds ratios and 95% CIs for diabetes and hypertension diagnosis. Pooled estimates were not estimated for CKD due to the heterogeneity of diagnostic measures between studies nor for cardiovascular disease due to the limited number of studies available. We assessed the variation between studies using the I^2^ statistic [29]. An Egger linear regression test and funnel plot was used to assess for publication bias [30]. Statistical significance of departure from the Egger test null hypothesis of no bias was guided by an alpha level of 0.05.

### Sensitivity Analyses

We conducted multiple sensitivity analyses. First, we stratified diabetes and hypertension summary estimates by age group (25-35, 35-45, 45+). Diabetes summary estimates were also stratified by diagnosis method. Additionally, to assess whether the implementation of the WHO Global NCD Action Plan in 2013 had an impact on screening and diagnosis of diabetes and hypertension, we conducted a sensitivity analysis excluding studies with enrollment periods prior to the Action Plan implementation in 2013.

All analyses were performed using STATA v. 15.

## Results

The database search yielded a total of 427 potentially relevant studies, of which 83 were duplicates. After screening 344 titles and abstracts, we excluded 303 studies, leaving 41 that met the criteria for full-text review (Figure 1). Of the 41 articles reviewed, 26 reported on a relevant outcome by ART status for at least one NCD of interest and were included in the final analysis. Some studies reported on multiple diseases of interest and are thus included in the analysis multiple times, once for each relevant outcome. A total of 16 studies reported on diabetes, 18 on hypertension, 6 on CKD, and one on CVD (Table 1). The total population of PLWH in our study was 13,570 [n = 8318 (61%) with reported ART use and n = 5252 (39%) with no reported ART use]. Sample size ranged from 104 to 3170 participants. Enrollment of cohorts began as early as 2008 and continued until at least 2016. Eight studies did not report enrollment dates. While a number of longitudinal cohort studies were included in the review, the primary outcomes measured longitudinally were not the outcomes of interest for this review. Outcomes of interest for this review, rather, were reported in Table 1 of the study. As such, all data included in this review is a cross-sectional report of outcomes of interest by ART status.

**Figure 1.**
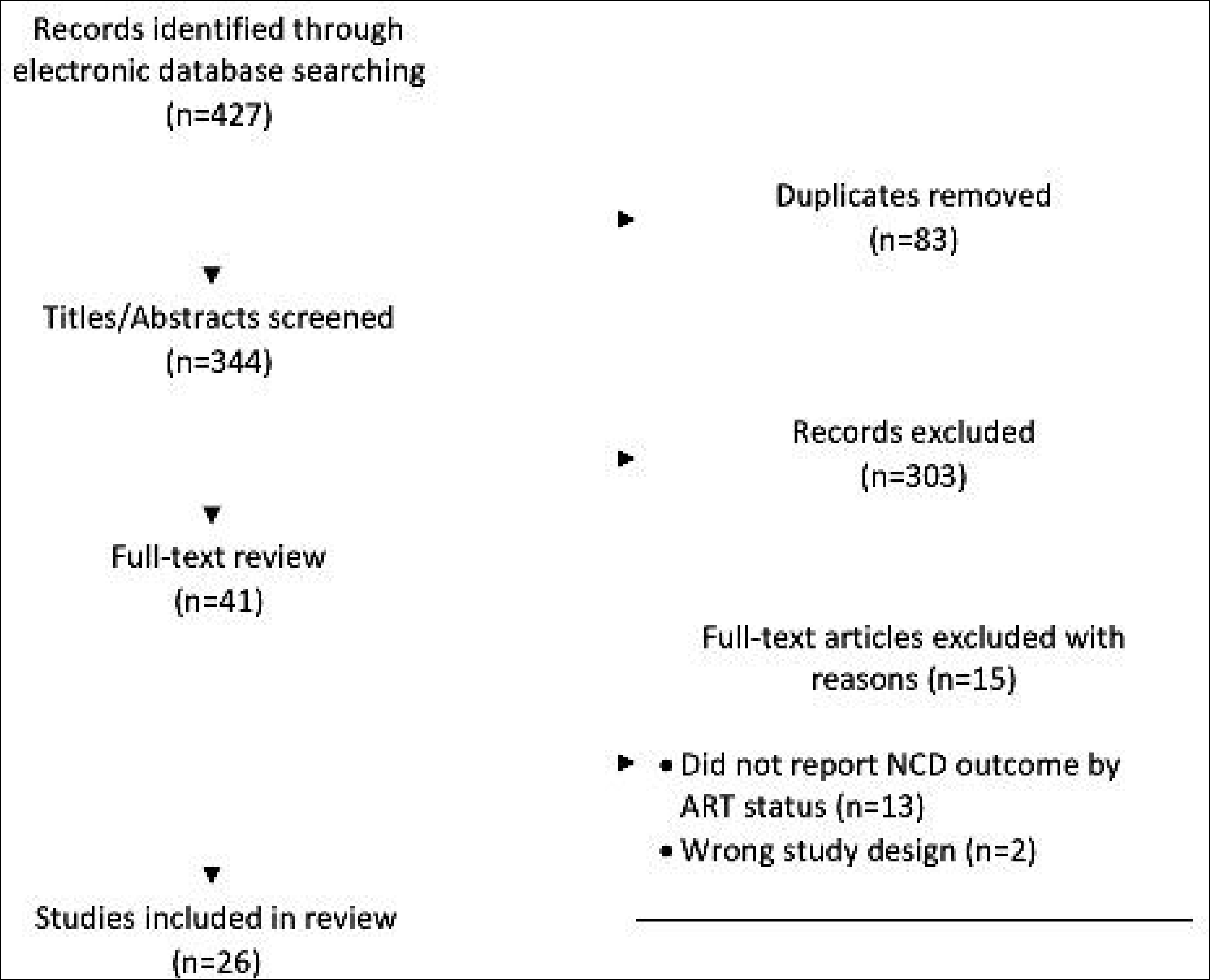
Flow chart of study selection

Similarly, ART use was not the primary exposure of interest in most studies included in this review, therefore detailed reporting on how ART status was assessed was not available in all studies. Of the studies that did report assessment of ART status, most relied upon self-report and/or medical records.

### Diabetes

Of the 16 studies addressing diabetes, only one reported *screening* prevalence by ART status (Table 1). Results of this study showed a slight increase in the likelihood of diabetes screening for PLWH with reported ART use (49%) compared to PLWH with no reported ART use (42%). The odds of diabetes *diagnosis* comparing PLWH with reported ART use to PLWH with no reported ART use ranged from 0.32 – 32.93 (Figure 2). Just over half of the studies showed increased odds of diabetes diagnosis among those with reported ART use. The pooled estimate of diabetes diagnosis showed that, among PLWH, reported ART use was associated with a 7% increase in the odds of being diagnosed with diabetes compared to no reported ART use (odds ratio (OR): 1.07; 95% CI: 0.71, 1.60). The confidence interval for this estimate, however, was imprecise and consistent with up to a 29% decrease and 60% increase in odds of diabetes diagnosis. The I^2^ statistic for the pooled diabetes diagnosis estimate was 62.1% (p-value=0.001). Only one study [21] reported on diabetes *treatment* by ART status and showed a greater likelihood of treatment initiation among PLWH with reported ART use (55%) compared to PLWH with no reported ART use (44%) (Table 1). No study assessed diabetes *control* among those who were treated.

**Figure 2.**
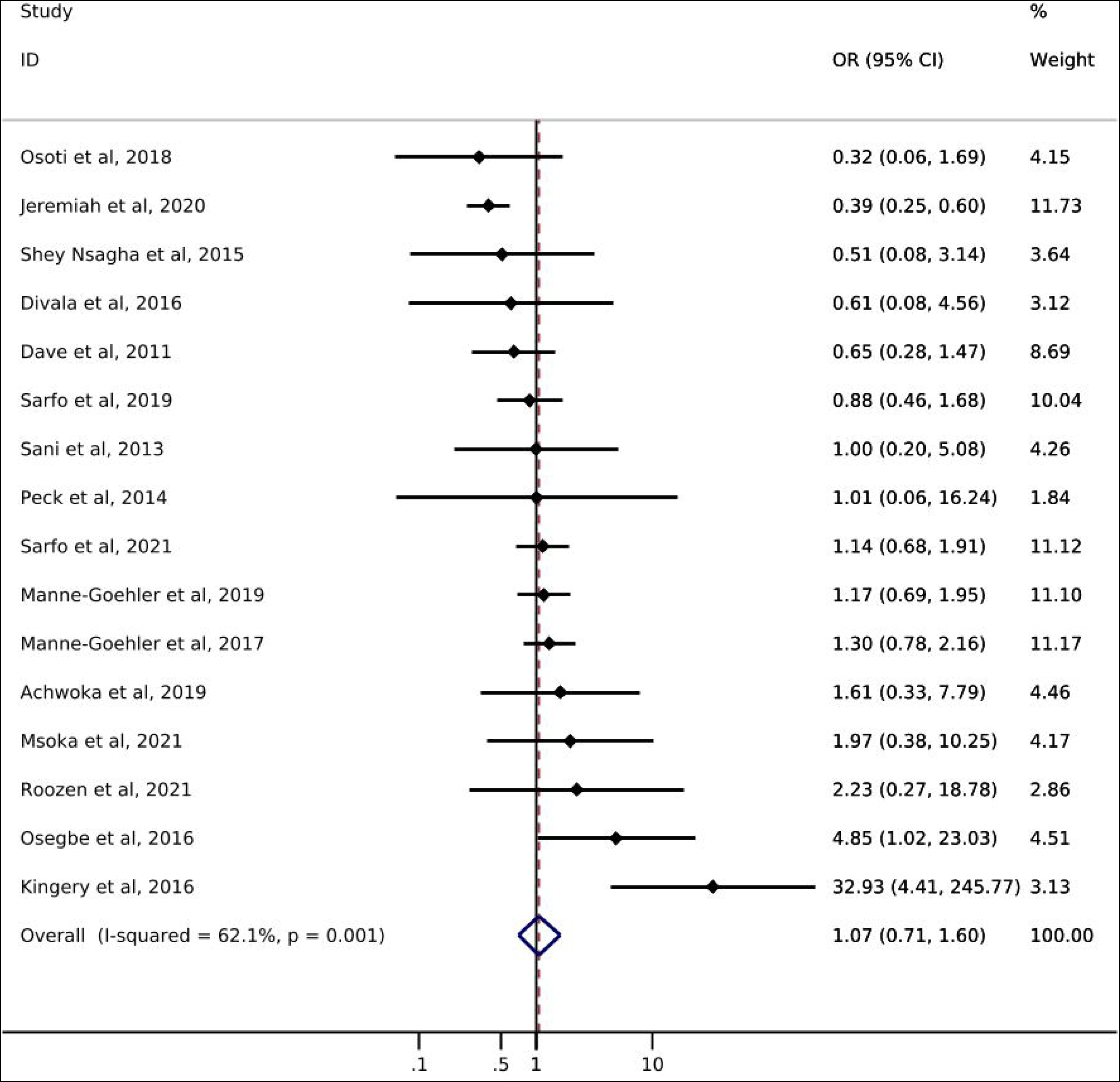
Forest plot of odds ratios for diabetes diagnosis comparing PLWH with reported ART use to PLWH with no reported ART use (n=16).

### Hypertension

Of the 18 studies reporting on hypertension, only one study from South Africa reported on hypertension *screening* by ART status. Manne-Goehler et al. [21] showed a slightly higher likelihood of hypertension screening for PLWH with reported ART use (72%) compared to PLWH with no reported ART use (69%) (Table 1). Odds of hypertension *diagnosis* ranged from 0.63 – 10.04 (Figure 3). All but three studies showed an increase in the odds of hypertension among PLWH with reported ART use compared to those without reported ART use. The pooled summary estimate showed a 110% increase in the odds of hypertension diagnosis among those with reported ART use compared to no reported ART use (OR: 2.10; 95% CI: 1.42, 3.09). The I^2^ statistic for the pooled hypertension diagnosis estimate was 87.4% (p-value=0.000). Five of the 18 studies reported on *treatment* of hypertension by ART status (Table 1). The study with the largest sample size (N=566) showed that PLWH with reported ART use were somewhat more likely to report use of anti-hypertensive medication than were those with no reported ART use (58% vs 49%). Results from the other four studies showed little difference between groups or had sample sizes too small to allow for meaningful comparisons. One study [31] reported hypertension *control* by reported ART status and showed that PLWH with reported ART use were less likely to achieve hypertension control (29.7%) than PLWH with no reported ART use (42.9%).

**Figure 3.**
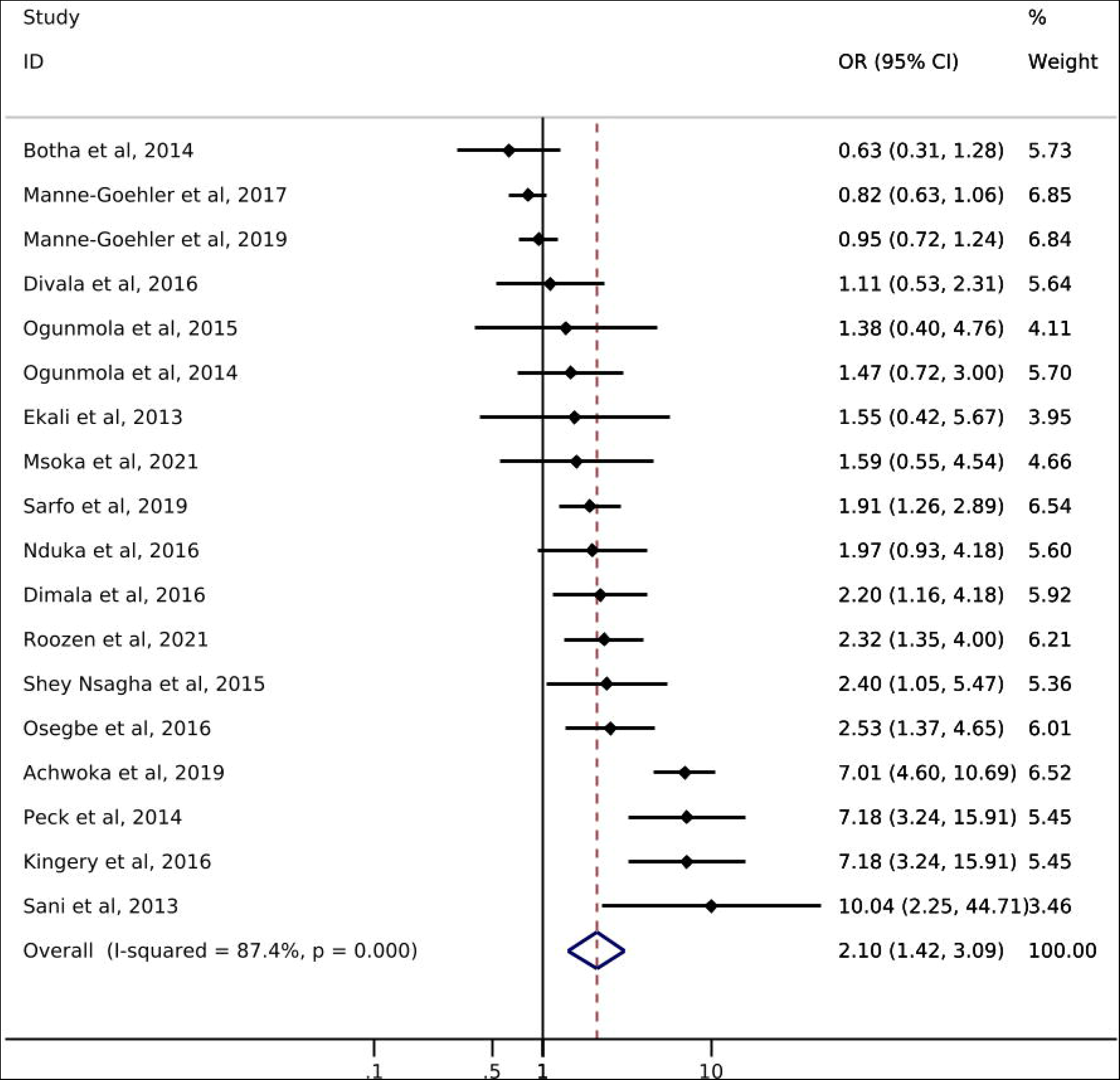
Forest plot of odds ratios for hypertension diagnosis comparing PLWH with reported ART use to PLWH with no reported ART use (n=18).

### Chronic Kidney Disease

Six studies reported on CKD diagnosis by ART status. The odds of CKD diagnosis when comparing those with reported ART use to those with no reported ART use ranged from 0.23 to 2.7 (Table 1). The odds of CKD diagnosis was higher in studies using creatine clearance to diagnose CKD, compared to studies basing CKD diagnosis according to estimated glomerular filtration rate (eGFR) (Table 1). No study reported on *screening* or *management* of CKD by ART status.

### Cardiovascular Disease

One study from Kenya reported on CVD by ART status among PLWH. Achwoka et al. [32] showed a higher prevalence of CVD (hypertension, heart attack, or stroke) among PLWH on ART (93.9%) compared to not on ART (53.9%). However, most identified cases of cardiovascular disease in this study were associated with elevated blood pressure. No studies reported on *screening*, *treatment* or *control* of CVD.

### Sensitivity analyses

### Age impact

The odds of diabetes diagnosis associated with ART use is generally similar across age groups (Supplemental Figure 1). In the age-stratified analysis of hypertension, the 35-45 age group showed higher odds of hypertension diagnosis (OR: 2.32 (95% CI: 1.72, 3.14)) compared to the 45+ age group (OR: 0.88 (95% CI: 0.73, 1.05)) (Supplemental Figure 2). Only one study reported a mean cohort age <35.

### Diagnostic test impact

When stratified by diagnosis method, studies using a glucose test had higher odds of diabetes diagnosis (OR: 1.51 (95% CI: 0.77, 2.96)) compared to studies using HbA1c (OR: 0.56, 95% CI: 0.26, 1.24) or self-report (OR: 1.12, 95% CI: 0.74, 1.70) (Supplemental Figure 3).

### WHO Global NCD Action Plan in 2013 impact

Finally, among studies enrolling participants after the implementation of the WHO Global NCD Action Plan in 2013 [33], the pooled estimate of effect for diabetes diagnosis showed an 18% decrease in the odds of diagnosis among PLWH with reported ART use compared to PLWH without reported ART use (OR: 0.82; 95% CI: 0.47, 1.41) (Supplemental Figure 4). The pooled estimate of effect for hypertension was slightly attenuated when estimated within this restricted study sample (OR: 1.80; 95% CI: 0.96, 3.41) (Supplemental Figure 5).

### Publication bias

Egger’s test to assess for publication bias yielded non-significant results for diabetes (p-value: 0.18) and hypertension (p-value: 0.05). Funnel plots showed minimal evidence of asymmetry suggesting little to no publication bias (Supplemental Figure 6).

### Quality Assessment

The possible range of quality scores was -8 to 16, with a higher score indicating better study quality. The overall quality of studies was high, with scores ranging from 9 – 16, and a median score of 12. The overall quality of studies may be higher than what is reported here as our analysis did not use the primary exposure/outcome measures for all studies.

## Discussion

This review summarizes available data on screening, diagnosis, treatment, and control of diabetes, hypertension, CKD, and CVD among PLWH on, vs not on, ART in sub-Saharan Africa. There has been a push to address the growing burden of NCDs through bolstering of existing healthcare systems for over a decade. In 2011, the Joint United Nations Program on HIV/AIDS (UNAIDS) recommended an integrated care model leveraging HIV treatment programs to scale up services for NCDs [34]. In 2013, the World Health Organization (WHO) followed suit with a similar recommendation in the Global Action Plan for the Prevention and Control of NCDs for 2013-2020 [35]. Moreover, many countries in SSA, including South Africa, have set up model clinics with integrated services [36]. Given these global and localized efforts, we hypothesized that among PLWH, being on ART--and thus engaged in HIV care--would result in higher rates of screening, diagnosis, treatment, and control of NCDs compared to not being on ART. However, our search over a ten-year timespan and across multiple databases yielded only 26 studies reporting on NCD diagnosis rates among a population of PLWH by ART status, and even fewer reports on screening, treatment, and control. The lack of data inhibits our ability to evaluate the full cascade of NCD care among PLWH and suggests limited update of NCD services, even among those who are engaged in ART care.

We found only one study over the last decade, conducted in South Africa, that reported on the full cascade of diabetes care (i.e., diabetes screening, diagnosis, and treatment rates) among a population of HIV-infected adults by ART status. Manne-Goehler et al [21] showed that among PLWH, those with reported ART use were slightly more likely to be screened, diagnosed and on treatment for diabetes, providing initial support to the notion that being on treatment for HIV may increase the rate of screening, diagnosis, and treatment of diabetes. The outcome of diabetes diagnosis was the only one that was complete for all 16 studies. In our meta-analysis of the 16 studies, we found a null association (OR: 1.07) between reported ART use and diabetes diagnosis. Though the range of study findings was vast (ORs ranged from 0.32 – 33) and most estimates were imprecise, 8 of the 16 studies included in the analysis reported an increased odds of diabetes diagnosis associated with reported ART use among PLWH. Without adequate data on diabetes screening rates among PLWH, we are unable to discern whether the higher odds of diabetes diagnosis seen in some of these studies is a result of being engaged in care, a side effect of ART use leading to higher diabetes prevalence, or a combination of the two. It is reasonable to postulate, however, that more frequent contact with a healthcare system would result in higher rates of diagnosis.

Our results showing increased odds of hypertension diagnosis among those with reported ART use may be due, in part, to ART-induced risk factors contributing to elevated blood pressure levels among PLWH [37],[38]. The ART care structure would seem to provide an ideal framework for treatment and monitoring of blood pressure levels in this higher-risk population [39]. Five studies reporting on hypertension treatment, however, showed very little difference in treatment rates by ART status, and the one study that reported on hypertension control among PLWH showed that those with reported ART use were less likely to achieve control. The lack of data on the full cascade of care inhibits our ability to truly understand the impact of HIV care programs on screening, diagnosis, treatment, and control of hypertension. However, the limited data in this review does suggest that HIV programs have not yet been successfully leveraged to improve hypertension prevention and control.

There is a well-established association between HIV and kidney disease [13],[40],[41]. Chronic inflammation as a result of HIV infection and high levels of toxicity due to long-term ART use can increase a person’s risk for acute kidney injury and CKD [13],[42],[41]. Consequently, we expected to see ample reports of kidney function monitoring efforts among PLWH and on ART. Yet, we found no studies in sub-Saharan African reporting on CKD screening rates, and very few reporting on CKD diagnosis rates, among PLWH and engaged in care. Given the established increase in risk, screening for CKD with creatinine clearance and urine dipsticks should be routinely conducted among PLWH, particularly among those on ART. The lack of data on CKD screening and diagnosis rates among PLWH in sub-Saharan Africa suggests a lack of monitoring in this population, despite a well-established care structure that would support frequent screening and early diagnosis of CKD.

We found only one study reporting on CVD diagnosis among a population of PLWH by ART use. Achwoka et al [32] showed a higher proportion of CVD among PLWH with reported ART use. However, the majority of cardiovascular events in this study population were related to elevated blood pressure which is more reflective of hypertension risk than CVD. Emerging data from high-income regions suggest that ART use contributes to an increased risk of CVD through alterations of lipid metabolism [43],[14]. The ART care structure provides a framework for critical monitoring of blood pressure and glucose levels, which would allow for early detection of risk factors of cardiovascular disease and mitigate progression and complications.

Our review should be interpreted in the context of its limitations. First, there is the possibility of incomplete retrieval or abstraction of data, or narrow search criteria resulting in missed studies. We did not obtain raw data from study investigators for pooled estimation and relied on simple proportions or estimates of association presented in their data. Diagnosis methods for both NCDs varied across studies, and many studies reporting hypertension diagnosis relied on self-report or medical record review, which could result non-differential outcome misclassification and likely underestimate the true association. Although heterogeneity between studies was high, we used random effects models to account for variability both within and between studies [44],[45]. However, the results may still represent a weighted average of a biased sample. We saw potential publication bias with regards to hypertension, however, given previous research showing higher rates of hypertension among PLWH, this may be related to a causal association rather than reporting bias. Due to lack of data on NCD care among PLWH, we were unable to evaluate the full cascade of care, including screening, treatment initiation, and control.

## Conclusion

The existing HIV care framework in SSA offers a promising setting to screen for NCDs among PLWH. A primary objective of the WHO’s 2013-2020 Global Action Plan was to not only strengthen existing health systems to address the burden of NCDs, but to also support high-quality research for the prevention and control of NCDs [46]. We conclude from this review, however, that evidence supporting that HIV care programs are successfully being leveraged to improve screening, diagnosis, treatment, and control of NCDs is lacking. Continued effort should be made to incorporate NCD services into HIV care programs in sub-Saharan Africa.

Furthermore, efforts to provide reporting on NCD screening, diagnosis, treatment, and control rates, would aid researchers, clinicians, and governments alike in understanding the true NCD risk among PLWH and the overall impact of care integration models.

## Data Availability

All data produced in the present study are contained in the manuscript

## Acknowledgements

Emma M Kileel: conception and study design, acquisition of data, data analysis, interpretation of data, drafting of work, final approval. Amy Zheng: acquisition of data, interpretation of data, revision of work, final approval. Jacob Bor: interpretation of data, revision of work, final approval. Matthew P Fox: interpretation of data, revision of work, final approval. Nigel J Crowther: interpretation of data, revision of work, final approval. Jaya A George: interpretation of data, revision of work, final approval. Siyabonga Khoza: interpretation of data, revision of work, final approval. Sydney Rosen: interpretation of data, revision of work, final approval. Willem DF Venter: interpretation of data, revision of work, final approval. Frederick Raal: interpretation of data, revision of work, final approval. Patricia Hibberd: interpretation of data, revision of work, final approval. Alana T Brennan: conception and study design, data analysis, interpretation of data, drafting of work, revision of work, final approval.

