## Supplemental Material for "Does engagement in HIV care affect screening, diagnosis, and control of noncommunicable diseases in sub-Saharan Africa? A systematic review and meta-analysis"

**Supplemental Table 1.** Systematic review search terms and databases

| *Electronic databases* | | | |
| --- | --- | --- | --- |
| **Database** | **Filters** | **Search** | **Results** |
| PubMed | 2011-2022 | (((((sub-saharan africa) AND (antiretroviral therapy)) AND ("diagnosis"[MeSH Subheading] OR "diagnosis"[All Fields] OR "screening"[All Fields] OR "mass screening"[MeSH Terms] OR ("mass"[All Fields] AND "screening"[All Fields]) OR "mass screening"[All Fields])))) AND ((diabetes mellitus) OR (kidney disease) OR (cardiovascular disease) OR (noncommunicable diseases)) | N=212 |
| Excerpta Medica Abstract Journals (EMBASE) | 2011-2022 | 'africa south of the sahara' AND 'antiretrovirus agent' AND ('screening' OR 'treatment' OR 'diagnosis') AND ('diabetes mellitus' OR 'kidney disease' OR 'cardiovascular disease' OR 'noncommunicable disease*') | N=19 |
| Web of Science (Science Citation Index and Social Science Citation Index) Includes Medline | 2011-2022 | ALL=(sub-saharan africa) AND ALL=(antiretroviral therapy ) AND ALL=(screening OR diagnosis OR treatment) AND ALL=(diabetes mellitus OR kidney disease OR cardiovascular disease OR noncommunicable diseases) | N=184 |
| *Conference websites* | | | |
| **Disease** | **Conference** | **Search** | **Results** |
| HIV | Conference on Retroviruses and Opportunistic Infections (CROI) | A. “Sub-Saharan Africa” AND “Antiretroviral Therapy” AND “Cardiovascular Disease” B. “Sub-Saharan Africa” AND “Antiretroviral Therapy” AND “Diabetes” C. “Sub-Saharan Africa” AND “Antiretroviral Therapy” AND “Kidney disease” D. “Sub-Saharan Africa” AND “Antiretroviral Therapy” AND “Noncommunicable diseases” | A.N=7  B.N=4*  C.N=0  D.N=1*  *Total N=9**  **3 duplicates in searches B and D from search A* |
|  | International AIDS Society (IAS) and AIDS Conference, site:  <https://www.abstract-archive.org> | A. “Sub-Saharan Africa” AND “Antiretroviral Therapy” AND “Cardiovascular Disease”* B. “Sub-Saharan Africa” AND “Antiretroviral Therapy” AND “Diabetes”* C. “Sub-Saharan Africa” AND “Antiretroviral Therapy” AND “Kidney disease”* D. “Sub-Saharan Africa” AND “Antiretroviral Therapy” AND “Noncommunicable diseases”*   **Due to search yield of 0, searches were rerun without “Antiretroviral Therapy”* *and all still yielded 0 results*   E. “Noncommunicable diseases” | A.N=0  B.N=0  C.N=0  D.N=0  E.N=3 |
| Diabetes | International Diabetes Federation World Diabetes Congress | - 2013 - “HIV” - 2015 - “HIV” - 2017 - *scientific programme not available* - 2019 - “HIV” | 2013 N=0  2015 N=0  2017 N=0  2019 N=0 |
|  | American Diabetes Association | - 2011 - “Antiretroviral Therapy” AND “Sub-Saharan Africa” - 2012 - “Antiretroviral Therapy” AND “Sub-Saharan Africa” - 2013 - “Antiretroviral Therapy” AND “Sub-Saharan Africa” - 2014 - “Antiretroviral Therapy” AND “Sub-Saharan Africa” - 2015 - “Antiretroviral Therapy” AND “Sub-Saharan Africa” - 2016 - “Antiretroviral Therapy” AND “Sub-Saharan Africa” - 2017 - “Antiretroviral Therapy” AND “Sub-Saharan Africa” - 2018 - “Antiretroviral Therapy” AND “Sub-Saharan Africa” - 2019 - “Antiretroviral Therapy” AND “Sub-Saharan Africa” - 2020 - “Antiretroviral Therapy” AND “Sub-Saharan Africa” - 2021 - “Antiretroviral Therapy” AND “Sub-Saharan Africa” | 2011 N=0  2012 N=0  2013 N=0  2014 N=0  2015 N=0  2016 N=0  2017 N=0  2018 N=0  2019 N=0  2020 N=0  2021 N=0 |
| Cardiovascular Disease | AHA Scientific Sessions | - 2015 - “HIV” - 2016 - “HIV” - 2017 - “HIV” - 2018 - “HIV” - 2019 - “HIV” - 2020 - “HIV” | 2015 N=0  2016 N=0  2017 N=0  2018 N=0  2019 N=0  2020 N=0 |
|  | Hypertension Scientific Sessions | - 2017 - “HIV” - 2019 - “HIV” - 2020 = “HIV” | 2017 N=0  2019 N=0  2020 N=0 |
| Kidney Disease | Kidney Week | - 2012 - “Antiretroviral therapy” - 2013 - “Antiretroviral therapy” - 2014 - “Antiretroviral therapy” - 2015 - “Antiretroviral therapy” - 2016 - “Antiretroviral therapy” - 2017 - “Antiretroviral therapy” - 2018 - “Antiretroviral therapy” - 2019 - “Antiretroviral therapy” - 2020 - “Antiretroviral therapy” | 2012 N=0  2013 N=0  2014 N=0  2015 N=0  2016 N=0  2017 N=0  2018 N=0  2019 N=0  2020 N=0 |

**Supplemental Table 2.** Inclusion/exclusion criteria for study selection

| **Inclusion criteria** | **Exclusion criteria** |
| --- | --- |
| Geographic range: sub-Saharan Africa  Topic: diabetes/kidney disease/hypertension/cardiovascular disease screening and care among an HIV positive population  Design: any  Population: adult patients with HIV (>=16 years)  Follow-up period: any  Time span of cohort: bo limit  Minimum information included: must report at least prevalence of screening, diagnosis, or treatment of at least one NCD of interest by ART status | Geographic range: outside of sub-Saharan Africa  Age <16  Pregnant women |

**Supplemental Table 3.** Study-defined definitions of noncommunicable disease screening, treatment, and control.

| **Study No** | **Study** | **Screening** | **Treatment** | **Control** |
| --- | --- | --- | --- | --- |
| **Outcome=Diabetes** | | | | |
| 8 | Manne-Goehler et al, 2017 | Answered ‘yes’ to survey question “Has a doctor, nurse, or other healthcare worker ever measured your urine or blood for diabetes?” | Answered ‘yes’ to either of the following: “Have you ever received treatment for diabetes prescribed by a doctor, nurse, or other healthcare worker?”, “Are you currently receiving any treatment for diabetes prescribed by a doctor, nurse, or other healthcare worker?” | - |
| **Outcome=Hypertension** | | | | |
| 18 | Botha et al, 2014 | - | Reported use of anti-hypertensive medication at time of interview | - |
| 7 | Kingery et al, 2016 | - | Reported use of antihypertensive treatment | - |
| 22 | Nduka et al, 2016 | - | Reported use of antihypertensive medication | - |
| 8 | Manne-Goehler et al, 2017 | Answered ‘yes’ to survey question “Has a doctor, nurse, or other healthcare worker ever measured your blood pressure?” | Answered ‘yes’ to “Are you currently on treatment for high blood pressure prescribed by a doctor, nurse, or other healthcare worker?” | - |
| 12 | Sarfo et al, 2019 | - | Reported use of antihypertensive medication | Blood pressure less than 140/90 mmHg |

*Studies not reporting on screening, treatment, or control of an NCD of interest are not included in this table. For a full list of NCD diagnosis definitions see Table 1.

**Supplemental Figure 1.** Forest plot of odds ratios for diabetes diagnosis comparing PLWH with reported ART use to PLWH with no reported ART use, stratified by mean age of study cohort (n=13).

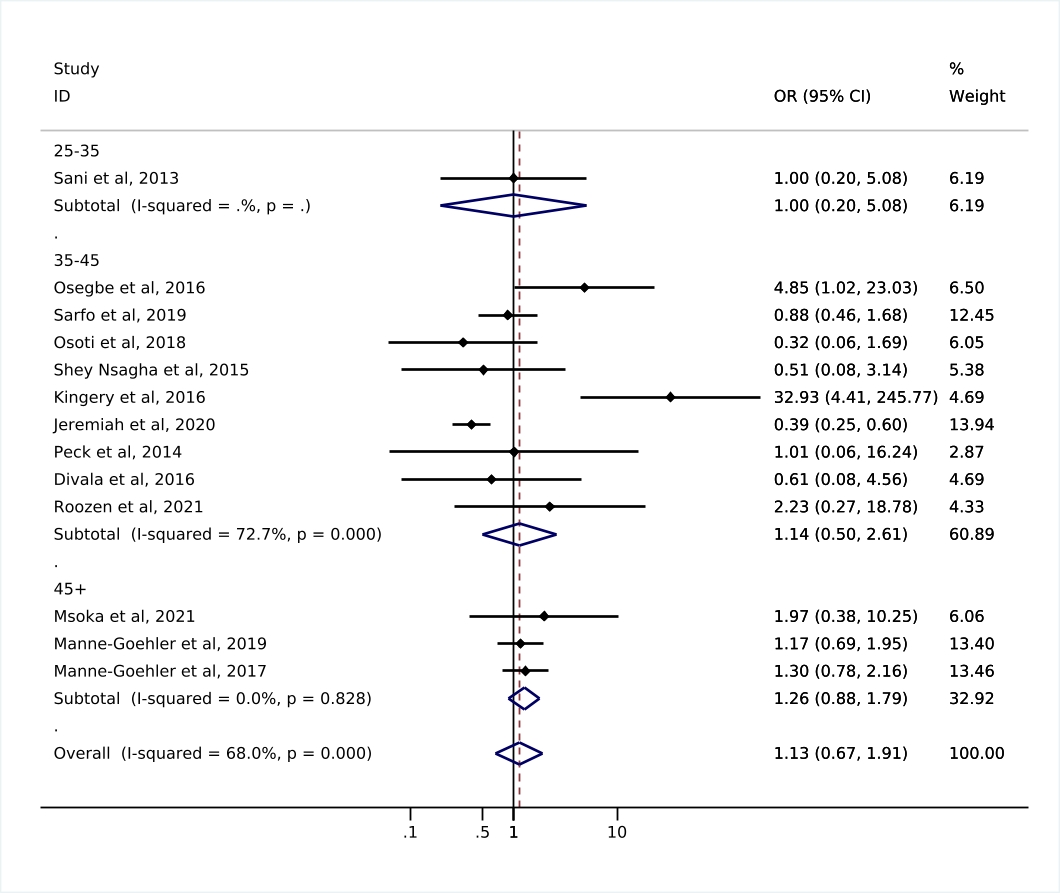

**Supplemental Figure 2.** Forest plot of odds ratios for hypertension diagnosis comparing PLWH with reported ART use to PLWH with no reported ART use, stratified by mean age of study cohort (n=17).

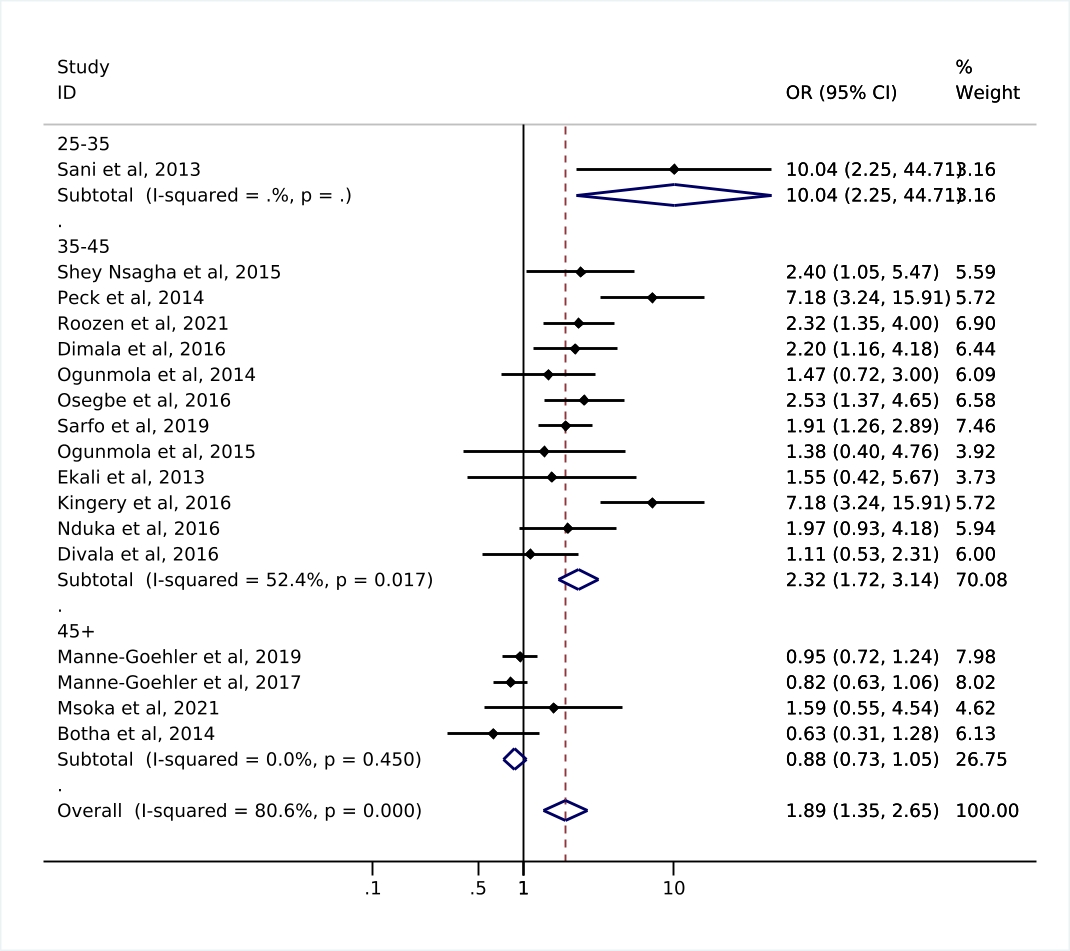

**Supplemental Figure 3.** Forest plot of odds ratios for diabetes diagnosis comparing PLWH with reported ART use to PLWH with no reported ART use, stratified by diagnosis method (n=15).

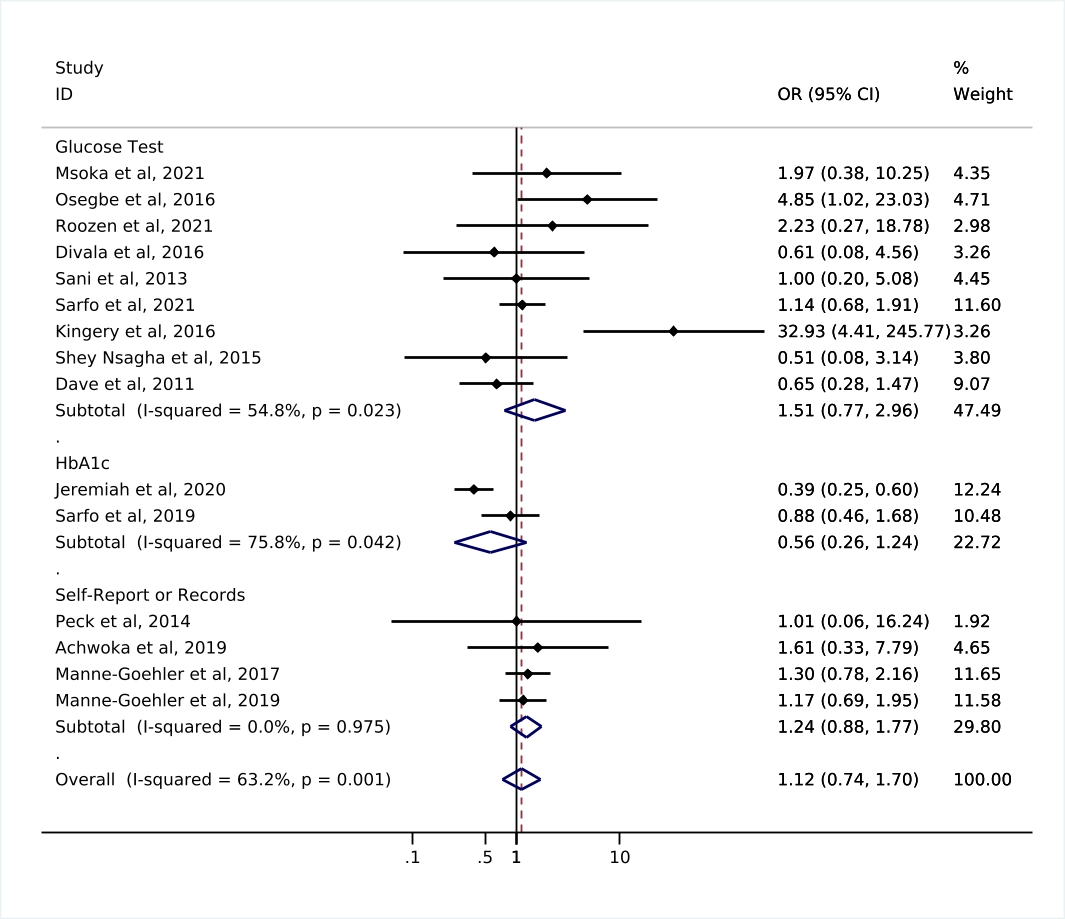

**Supplemental Figure 4.** Forest plot of odds ratios for diabetes diagnosis comparing PLWH with reported ART use to PLWH with no reported ART use, restricted to studies enrolling participants after the implementation of the WHO Global NCD Action Plan in 2013 (n=8).

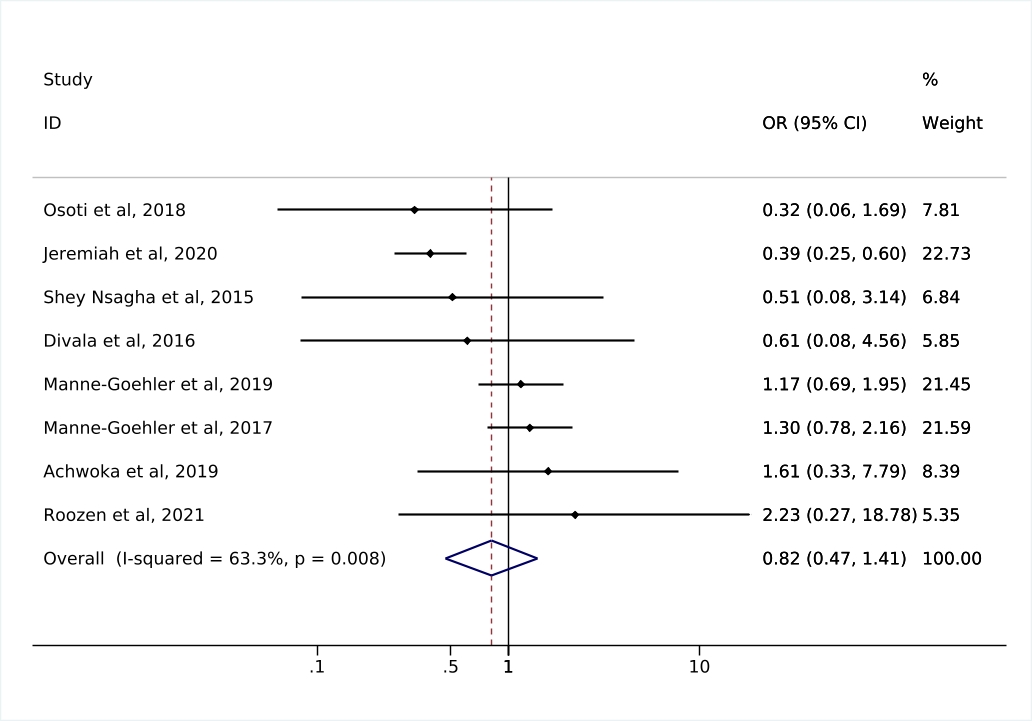

**Supplemental Figure 5.** Forest plot of odds ratios for hypertension diagnosis comparing PLWH with reported ART use to PLWH with no reported ART use, restricted to studies enrolling participants after the implementation of the WHO Global NCD Action Plan in 2013 (n=7).

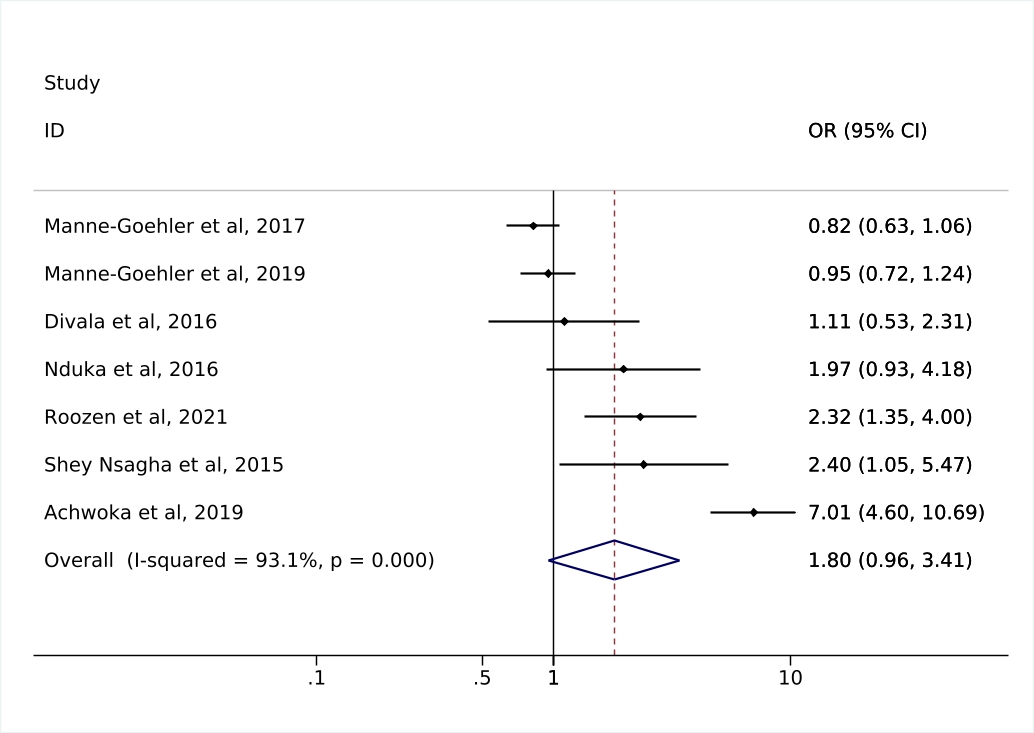

**Supplemental Figure 6.** Funnel plots assessing publication bias

| A) | B) |
| --- | --- |
| 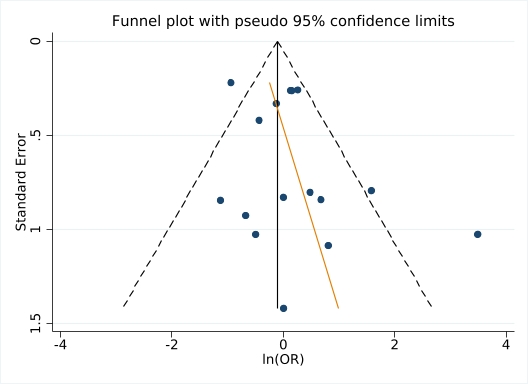 | 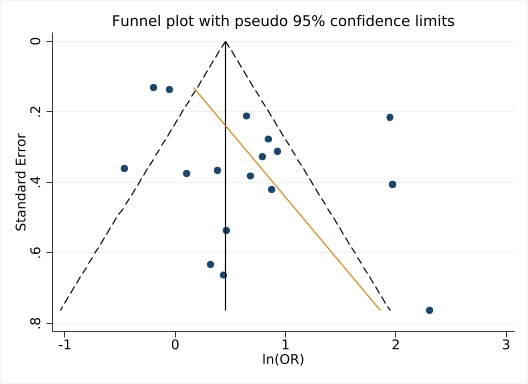 |

**Funnel plot assesses the hypothesis that the relationship between log(OR) and standard error of the log(OR) are independent.

**Panel A: funnel plot assessing publication bias of *diabetes diagnosis.* Egger’s test p-value=0.18.

**Panel B: funnel plot assessing publication bias of *hypertension diagnosis.* Egger’s test p-value=0.054.
